# Congruence between preferred and actual place of death and its association with patients’ quality of death and dying: A nationwide survey in Japan

**DOI:** 10.1101/2025.02.22.25322724

**Authors:** Mariko Shutoh, Tatsuya Morita, Maho Aoyama, Yoshiyuki Kizawa, Yasuo Shima, Mitsunori Miyashita

## Abstract

**Background:** Satisfying patients’ preferences is an important outcome in palliative care. Previous research has reported that a patient’s place of death was associated with quality of death and dying.

**Purpose:** This study aimed to assess factors associated with congruence between a patient’s preferred and actual place of death and their quality of death and dying, as perceived by their family caregivers.

**Method:** Data were obtained from a nationwide cross-sectional questionnaire survey of bereaved family caregivers of patients with cancer in Japan. A total of 13,711 family caregivers participated. We evaluated the quality of death and dying using the Good Death Inventory.

**Results:** Patients who died in their preferred place were categorized as the “achieved group,” whereas patients who died in a place that they did not prefer were classified as the “not-achieved group.” Survey scores were significantly higher for the achieved group compared with the not-achieved group (48.8±10.1 and 44.0±9.5, respectively; p <0.001). A multiple linear regression analysis indicated that congruence between preferred and actual place of death was an independent determinant of good quality of death and dying (p <0.001).

**Conclusion:** Congruence between a patient’s preferred and actual place of death may contribute to better quality of death and dying among terminally ill patients with cancer. Congruence between preferred and actual place of death should be regarded as an essential goal in end-of-life care.

## Introduction

In Japan, approximately 370,000 people die of cancer each year. Of these, only 11.8% die at home, 16.5% die in a palliative care unit (PCU), and 68.1% die in hospital [1].

Receiving care and dying in one’s preferred place is one of the essential factors for a high quality of death and dying [2–3].

More than half of surveyed populations would prefer to be cared for and die at home, and the quality of death and dying is deemed better at home than in hospital [4–6]. Additionally, more than half of patients with terminal cancer prefer home hospice care but most die in an institution. The reasons are complex, with various determinants influencing decisions regarding where end-of-life care is provided [7–10].

To our knowledge, the effects of congruence between preferences for and actual place of death on quality of patient death have not been elucidated sufficiently. The aim of this study was to evaluate the association between this congruence and patients’ quality of death and dying, as perceived by their family caregivers. We hypothesized that patients who died at their preferred place had better end-of-life quality compared with the patients who were not able to be in their preferred place when they died.

## Methods

This was a nationwide, cross-sectional, anonymous, self-report questionnaire survey for bereaved family members of deceased cancer patients. This study was one part of the Japan Hospice and Palliative Care Evaluation Study 3 (J-HOPE3). The detailed methodology used in this survey has been described previously [11].

### Participating institutions

We sent letters to 396 institutions that, prior to July 1, 2013, were members of Hospice Palliative Care Japan (HPCJ), an organization of palliative care institutions in Japan. Of these institutions, 49 were acute-care hospitals, 296 were inpatient PCUs, and 51 were home hospice services. We received valid responses from 175 institutions comprising 20 acute hospitals, 133 PCUs, and 22 home hospice services.

### Participants

We conducted a cross-sectional, anonymous, self-report questionnaire between May and July 2014. To identify potential patients, we asked each institution to identify and list up to 80 bereaved family members of patients who had died prior to October 2012. The inclusion criteria were: (1) the patient died of cancer; (2) the patient was at least 20 years old; and (3) the bereaved family member was at least 20 years old. The exclusion criteria were: (1) the patient received palliative care for less than 3 days; (2) the bereaved family member could not be identified; (3) treatment-associated death or death occurred in an intensive care unit; (4) the potential participant was at risk of suffering serious psychological distress, as determined by the primary physician and a nurse; and (5) the potential participant was incapable of completing the self-report questionnaire because of cognitive impairment or visual disability. The questionnaire was sent to the bereaved family members from each participating institution along with a letter explaining the survey. The return of a completed questionnaire was considered consent to participate in the study. A ballpoint pen was included in the envelope as an incentive to participate. Participants were asked to return their completed questionnaires to the secretariat office (Tohoku University) within 2 weeks. We sent a reminder to non-responders 1 month after sending the questionnaire. If they did not wish to participate in the study, they were asked to check a “no participation” box and return the incomplete questionnaire. Ethical approval for the study was granted by the institutional review boards of Tohoku University and all participating institutions.

### Questionnaires

#### Preferred place for end-of-life care and death

Congruence between preferred and actual place of death was retrospectively assessed by asking family caregivers, “In which place of care did he or she wish to spend his or her last days?”

Participants were asked to choose a preferred place for end-of-life care and death from six options: a home setting, an acute-care hospital unit, a palliative care unit, others (e.g., nursing home), no preference, and preference unknown. We adopted this question on the basis of previous literature and expert opinions [4,12–14].

Patients who died in their preferred place were categorized as the “achieved group.” Patients who died in a place that was not their preference were classified as the “not-achieved group.”

### Good Death Inventory-short version

We used the short version of the Good Death Inventory (GDI) to measure patients’ achievement of a good death from the perspective of their bereaved family members. This measure was developed on the basis of qualitative interviews and a large-scale quantitative study, it has 18 domains representing concepts important to good death for Japanese patients with cancer. In this study, we used the short version of the GDI, which consists of 10 core items and has sufficient reliability and validity [15,16]. The caregiver was asked to rate each item using 7-point Likert scale (ranging from1: totally disagree to 7: absolutely agree). Total scores were calculated by summing the scores for all items, with a high total score indicating the achievement of a good death. Higher scores indicated a better quality of death. The GDI has been validated outside Japan [17].

### Covariates

Bereaved family caregivers were asked to report potential confounding factors that might be associated with the congruence between preferred and actual place of death and patients’ quality of death, for example: (1) Presence of other caregivers; (2) Communication with the patient about the disease and about life; and (3) Implementation of end-of-life discussions. We adopted these covariates on the basis of previous literature and expert opinions [4,12–14].

### Analysis

To define the potential determinants of congruence between preferred and actual place of death for patients with cancer, the dependent variables were split into two categories: the achieved group and the not-achieved group. The differences between the groups were determined using Chi-square tests.

*T*-test and Analysis of Variance (ANOVA) models were used to examine relationships between the congruence between patients’ preferred and actual place of death and their quality of death. Where significant differences were observed, a post hoc Dunnett test was used to explore between which pair of groups the differences lay.

We conducted linear regression analyses to evaluate associations between place of death and quality of death and dying.

All statistical analyses were performed with EZR (Saitama Medical Center, Jichi Medical University, Saitama, Japan), which is a graphical user interface for R (The R Foundation for Statistical Computing, Vienna, Austria). More precisely, it is a modified version of R commander designed to add statistical functions frequently used in biostatistics [18].

## Results

Invitations were sent to 396 institutions (49 acute-care hospitals, 296 PCUs, and 51 home hospice services). Of these, 20 acute-care hospitals, 133 PCUs, and 22 home hospice services participated.

We identified 15,632 potential participants, of which 1,921 were excluded for not meeting the inclusion criteria or for meeting the exclusion criteria. Thus, questionnaires were sent to the remaining 13,711 bereaved family members; 9,123 responses were finally analyzed (effective response rate was 67%). The data analysis included 814, 7292, and 1017 family members’ questionnaires from acute hospitals, PCUs, and home hospice services, respectively. The mean duration between patients’ death and completion of the questionnaire was 283.4 days (Standard deviation 139.5).

### Characteristics of patients and bereaved family members

The characteristics of the participating patients and bereaved family members are shown in Table 1. The age of most of the patients was ≥ 60 years, more than half were men, and the most common primary cancer site was the lungs or gastrointestinal tract.

**Table 1.** Characteristics of Patients and Family Caregivers.

| Patient characteristics | Total |  | Achieved |  | Not achieved |  | p value* |
| --- | --- | --- | --- | --- | --- | --- | --- |
|  | n | (%) | n | (%) | n | (%) |  |
| Number of people | 9123 | 100 | 3640 | 39.9 | 3487 | 38.2 |  |
| Gender |  |  |  |  |  |  | 0.59 |
| Man | 5094 | 55.8 | 2018 | 55.4 | 1979 | 56.8 |  |
| Age<br>Average : 73.5 SD : 11.5 |  |  |  |  |  |  | p < 0.001 |
| <40y | 69 | 0.8 | 28 | 0.8 | 24 | 0.7 |  |
| 40-49y | 228 | 2.5 | 84 | 2.3 | 94 | 2.7 |  |
| 50-59y | 681 | 7.5 | 276 | 7.6 | 275 | 7.9 |  |
| 60-69y | 1957 | 21.5 | 910 | 25.0 | 736 | 21.1 |  |
| 70-79y | 2747 | 30.1 | 1188 | 32.6 | 1061 | 30.4 |  |
| 80-89y | 2590 | 28.4 | 981 | 27.0 | 1113 | 31.9 |  |
| $\geq 90y$ | 490 | 5.4 | 163 | 4.5 | 219 | 6.3 | |
| Primary tumor site |  |  |  |  |  |  | 0.35 |
| Lung | 2103 | 23.1 | 815 | 22.4 | 826 | 23.7 |  |
| Digestive system | 4167 | 45.7 | 1722 | 47.3 | 1611 | 46.2 |  |
| Other | 2839 | 31.1 | 1096 | 30.1 | 1084 | 31.1 |  |
| Actual place of death |  |  |  |  |  |  | p<0.001 |
| Home | 1017 | 11.1 | 864 | 23.7 | 69 | 2.0 |  |
| Acute hospitals | 814 | 8.9 | 160 | 4.4 | 420 | 12.0 |  |
| PCU | 7292 | 79.9 | 2616 | 71.9 | 3036 | 87.1 |  |
| Annual Income during care (JPY) |  |  |  |  |  |  | 0.003 |
| < 2,000,000 | 2659 | 29.1 | 982 | 27.0 | 1111 | 31.9 |  |
| 2,000,000-3,999,999 | 3284 | 36.0 | 1369 | 37.6 | 1307 | 37.5 |  |
| ≥4,000,000 | 2444 | 26.8 | 1026 | 28.2 | 956 | 27.4 |  |
| Medical bills |  |  |  |  |  |  | 0.38 |
| < 100,000 | 2425 | 26.6 | 981 | 27.0 | 956 | 27.4 |  |
| 100,000-199,999 | 2869 | 31.4 | 1131 | 31.1 | 1153 | 33.1 |  |
| ≥200,000 | 3255 | 35.7 | 1364 | 37.5 | 1284 | 36.8 |  |
| Intervention of Palliative care team |  |  |  |  |  |  | 0.001 |
| Yes | 6718 | 73.6 | 2838 | 78.0 | 2644 | 75.8 |  |
| No | 1362 | 14.9 | 489 | 13.4 | 557 | 16.0 |  |
| unkown | 632 | 6.9 | 223 | 6.1 | 242 | 6.9 |  |
| Implementation of EOL discussion |  |  |  |  |  |  | 0.31 |
| Yes | 7352 | 80.6 | 3013 | 82.8 | 2955 | 84.7 |  |
| No | 1266 | 13.9 | 530 | 14.6 | 482 | 13.8 |  |
| Family Caregiver Characteristics |  |  |  |  |  |  |  |
| Number of people | 9123 | 100.0 | 3640 | 39.9 | 3487 | 38.2 |  |
| Gender |  |  |  |  |  |  | 0.45 |
| Male | 2940 | 32.2 | 1137 | 31.2 | 1236 | 35.4 |  |
| Age Average : 73.5 SD : 11.5 |  |  |  |  |  |  | p<0.001 |
| <40y | 367 | 4.0 | 117 | 3.2 | 159 | 4.6 |  |
| 40-49y | 1106 | 12.1 | 366 | 10.1 | 488 | 14.0 |  |
| 50-59y | 2211 | 24.2 | 780 | 21.4 | 915 | 26.2 |  |
| 60-69y | 2669 | 29.3 | 1132 | 31.1 | 1035 | 29.7 |  |
| 70-79y | 1893 | 20.7 | 899 | 24.7 | 684 | 19.6 |  |
| 80-89y | 575 | 6.3 | 283 | 7.8 | 197 | 5.6 |  |
| ≥90y | 17 | 0.2 | 10 | 0.3 | 5 | 0.1 |  |

| Relationship |  |  |  |  |  |  | p < 0.001 |
| --- | --- | --- | --- | --- | --- | --- | --- |
| Spouse | 4106 | 45.0 | 1589 | 43.7 | 1821 | 52.2 |  |
| Child | 3232 | 35.4 | 1383 | 38.0 | 1116 | 32.0 |  |
| Son-/daughter-in-law, Parent, Sibling, other | 1527 |  | 630 | 17.3 | 550 | 15.8 |  |
| Education |  |  |  |  |  |  | 0.54 |
| Elementary school to high school | 4984 | 22.2 | 2024 | 55.6 | 1989 | 57.0 |  |
| Vocational school, Junior college, Undergraduate, Graduate | 3812 | 17.0 | 1555 | 42.7 | 1482 | 42.5 |  |
| Presence of other caregivers |  |  |  |  |  |  | 0.30 |
| Present | 6433 | 70.5 | 2588 | 71.1 | 2588 | 74.2 |  |
| Absent | 2362 | 25.9 | 979 | 26.9 | 892 | 25.6 |  |
| Communication about the disease and life with the patient |  |  |  |  |  |  | 0.14 |
| Frequently | 3508 | 38.5 | 1725 | 47.4 | 1292 | 37.1 |  |
| As needed | 4567 | 50.1 | 1660 | 45.6 | 1897 | 54.4 |  |
| Rarely | 792 | 8.7 | 225 | 6.2 | 312 | 8.9 |  |

\*Differences between the groups were tested using a $\chi^2$ test.
PCU: Palliative Care Unit
Total of some items do not add to 100% owing to missing data.

The age of most of the bereaved family members was ≥ 50 years, nearly two-thirds were women, most were the spouse or child of the patient, and most had completed high school or higher education.

### Preferred place of death

The preferred places of death are shown in Table 2. Of the 9,123 patients in this study, 3,841 (42.1%) preferred their home for their place of death, 2,754 (30.2%) preferred a PCU, and 520 (5.7%) preferred an acute-care hospital. The congruence rates between preferred and actual place of death were 22.5%, 94.5%, and 30.8% for home, PCU, and acute-care hospital, respectively.

**Table 2.** GDI Scores According to Congruence between Preferred and Actual Place of Death.

|  | Total |  |  |  |  | home |  |  |  |  | hospital |  |  |  |  | PCU |  |  |  |  |
| --- | --- | --- | --- | --- | --- | --- | --- | --- | --- | --- | --- | --- | --- | --- | --- | --- | --- | --- | --- | --- |
|  | Achieved<br>n=3640 |  | Not achieved<br>n=3525 |  |  | Achieved<br>n=864 |  | Not achieved<br>n=69 |  |  | Achieved<br>n=160 |  | Not achieved<br>n=420 |  |  | Achieved<br>n=2616 |  | Not achieved<br>n=3036 |  |  |
|  | Ave | SD | Ave | SD | P value | Ave | SD | Ave | SD | P value | Ave | SD | Ave | SD | P value | Ave | SD | Ave | SD | P value |
| GDI | 48.8 | 10.1 | 44.0 | 9.5 | p<0.001 | 49.0 | 11.8 | 48.2 | 9.6 | 0.55 | 43.6 | 12.3 | 41.2 | 11.5 | 0.034 | 49.1 | 9.2 | 44.2 | 9.1 | p<0.001 |
| Physical ond psychological comfort | 5.3 | 1.4 | 4.9 | 1.5 | p<0.001 | 5.1 | 1.5 | 5.0 | 1.6 | 0.76 | 4.5 | 1.6 | 4.3 | 1.6 | 0.13 | 5.3 | 1.3 | 5.1 | 1.4 | p<0.001 |
| Dying in a favorite place | 5.7 | 1.3 | 4.1 | 1.6 | p<0.001 | 6.2 | 1.2 | 5.7 | 1.3 | p<0.001 | 4.9 | 1.3 | 4.0 | 1.8 | p<0.001 | 5.6 | 1.2 | 4.2 | 1.6 | p<0.001 |
| Maintaing hope and pleasure | 4.6 | 1.6 | 3.9 | 1.6 | p<0.001 | 4.9 | 1.5 | 4.8 | 1.7 | 0.42 | 3.9 | 1.6 | 3.6 | 1.7 | 0.088 | 4.6 | 1.6 | 3.9 | 1.6 | p<0.001 |
| Good relationship with medical staff | 5.8 | 1.1 | 5.3 | 1.3 | p<0.001 | 5.8 | 1.2 | 5.9 | 1.0 | 0.87 | 5.6 | 1.2 | 5.3 | 1.3 | 0.014 | 5.8 | 1.1 | 5.3 | 1.3 | p<0.001 |
| Not being a burden to others | 3.5 | 1.6 | 3.5 | 1.6 | 0.86 | 3.3 | 1.6 | 3.5 | 1.6 | 0.17 | 3.6 | 1.6 | 3.4 | 1.5 | 0.37 | 3.5 | 1.6 | 3.5 | 1.6 | 0.27 |
| Good relationship with family | 5.3 | 1.4 | 4.8 | 1.5 | p<0.001 | 5.5 | 1.3 | 5.0 | 1.5 | 0.002 | 4.9 | 1.5 | 4.5 | 1.5 | 0.010 | 5.2 | 1.4 | 4.8 | 1.4 | p<0.001 |
| Independence | 3.4 | 1.9 | 2.9 | 1.9 | p<0.001 | 3.4 | 1.9 | 3.1 | 1.8 | 0.20 | 3.6 | 1.9 | 3.5 | 1.9 | 0.71 | 3.4 | 1.9 | 2.8 | 1.8 | p<0.001 |
| Environmental comfort | 5.6 | 1.1 | 5.1 | 1.3 | p<0.001 | 5.7 | 1.2 | 5.4 | 1.1 | 0.076 | 5.0 | 1.3 | 4.6 | 1.5 | 0.002 | 5.6 | 1.1 | 5.2 | 1.3 | p<0.001 |
| Being respected as an individual | 6.1 | 0.9 | 5.9 | 1.1 | p<0.001 | 6.2 | 0.9 | 6.0 | 1.0 | 0.092 | 5.7 | 1.2 | 5.6 | 1.2 | 0.33 | 6.1 | 0.9 | 5.9 | 1.1 | p<0.001 |
| Life completion | 4.9 | 1.8 | 4.4 | 1.8 | p<0.001 | 4.9 | 1.8 | 5.0 | 1.6 | 0.710 | 4.4 | 1.8 | 4.1 | 1.8 | 0.12 | 4.9 | 1.7 | 4.5 | 1.8 | p<0.001 |

GDI: Good Death Inventory
PCU: Palliative Care Unit
Average: Ave

Of the 9,123 family caregivers, 2,388 (26.2%) preferred home as the place of death for their loved one, 5,038 (55.2%) preferred a PCU, and 636 (7.0%) preferred an acute-care hospital. The congruence rates between caregivers’ preferred and actual place of death for their loved ones were 32.1%, 93.2%, and 38.7% for home, PCU, and acute-care hospital, respectively.

### Congruence between preferred and actual place of death and its association with quality of death and dying

The mean GDI scores were 50.8±7.9, 46.9±8.4, and 43.5±9.1 for home, PCU, and acute-care hospital deaths, respectively.

Fig 1 and Table 2 show the mean GDI scores for both the achieved group and the not-achieved group.

**Fig 1.**
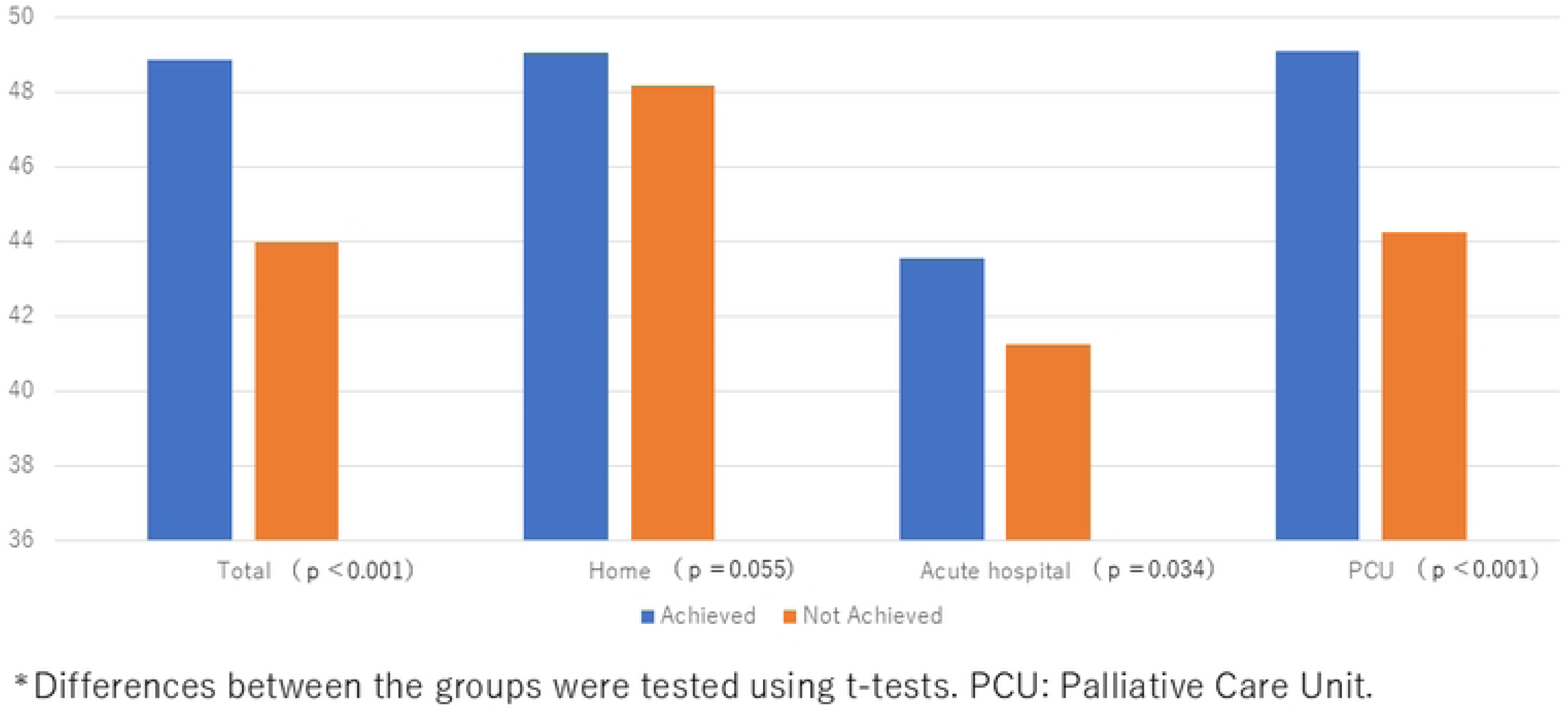
GDI Scores According to Congruence between Preferred and Actual Place of Death. *Differences between the groups were tested using t-tests. PCU: Palliative Care Unit.

With respect to the congruence between preferred and actual place of death, GDI scores were significantly higher for the achieved group than the not-achieved group (48.8±10.1 and 44.0±9.5, respectively; p <0.001).

For those who died in acute-care hospitals or PCUs, GDI scores were significantly higher among the achieved group than the not-achieved group. For those who died at home, nonsignificant differences were observed in GDI scores between groups.

A significant difference between groups was observed (achieved, not-achieved, no preference, and preference unknown), as determined by a one-way ANOVA (p <0.001). A post hoc Dunnett test showed that GDI scores were significantly higher for achieved compared with not-achieved, no preference, and preference unknown for preferred place of death (see S1 Table).

### Domains of Good Death Inventory according to congruence between preferred and actual place of death

All domains, aside from “not being a burden to others,” scored higher in the achieved group compared with the not-achieved group. Domain scores according to care setting are summarized in Fig 2 and S1–3 Figs.

**Fig 2.**
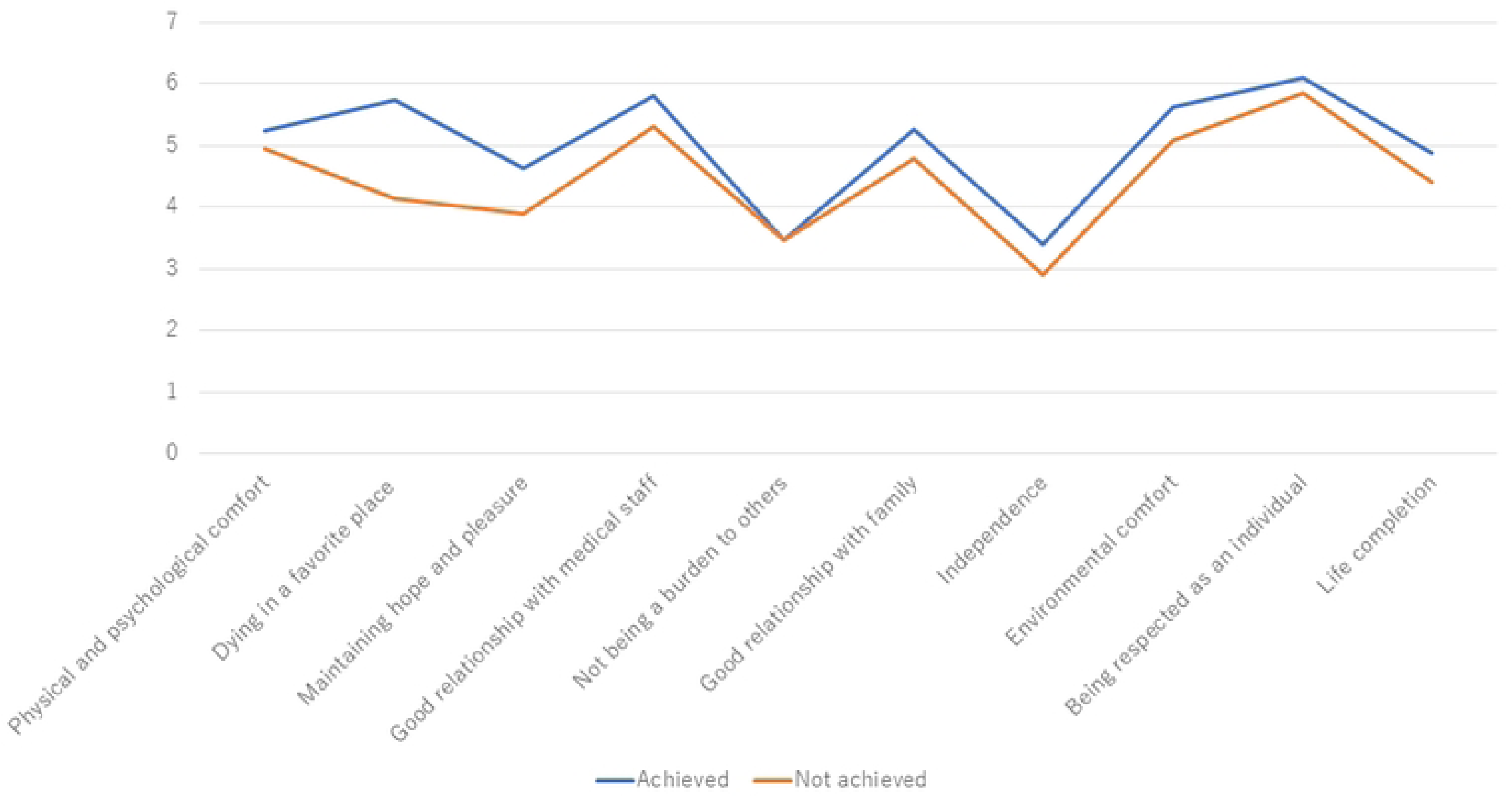
GDI Domain Scores According to Congruence between Preferred and Actual Place of Death.

### Factors related to quality of death and dying: Results of a multiple linear regression analysis

Table 3 shows the results of the linear regression analyses for the association between GDI scores and characteristics of patients and their families. GDI scores differed significantly as a function of patients’ gender, age, actual place of death, congruence between preferred and actual place of death, medical bills, intervention of a palliative team, presence of other caregivers, communication with the patient about their disease and life (all p <0.001).

**Table 3.** Factors Related to Quality of Death: Results of a Multiple Linear Regression Analysis.

|  | Regression<br>coefficient | 95%CI | P value |
| --- | --- | --- | --- |
| Patient characteristics |  |  |  |
| Gender |  |  |  |
| Man (reference) | 0 |  |  |
| Woman | -0.38 | -0.90 to 0.15 | 0.16 |
| Age | 0.09 | 0.06 to 0.12 | $p < 0.001$ |
| Primary tumor site |  |  |  |
| Lung (reference) | 0 |  |  |
| Digestive system | 0.46 | -0.17 to 1.04 | 0.12 |
| Other | 0.38 | -0.25 to 1.02 | 0.24 |
| Actual place of death |  |  |  |
| Home (reference) | 0 |  |  |
| Acute hospitals | -4.27 | -5.36 to -3.18 | p < 0.001 |
| PCU | -0.90 | -1.61 to -0.18 | 0.014 |
| Congruence between preferred and actual place of death |  |  |  |
| Achieved (reference) | 0 |  |  |
| Not achieved | -4.23 | -4.72 to -3.75 | p < 0.001 |
| Annual Income during care (JPY) |  |  |  |
| < 2,000,000 (reference) | 0 |  |  |
| 2,000,000-3,999,999 | 0.26 | -0.30 to 0.81 | 0.37 |
| ≥4,000,000 | 0.77 | 0.16 to 1.37 | 0.013 |
| Medical bills |  |  |  |
| < 100,000 (reference) | 0 |  |  |
| 100,000-199,999 | -0.26 | -0.47 to -0.05 | 0.095 |
| ≥200,000 | -1.00 | -1.59 to -0.42 | p < 0.001 |
| Intervention of Palliative care team |  |  |  |
| Yes (reference) | 0 |  |  |
| No | -1.05 | -1.69 to -0.41 | 0.001 |
| unkown | -2.21 | -3.16 to -1.25 | p < 0.001 |
| Implementation of EOL discussion |  |  |  |
| Yes (reference) | 0 |  |  |
| No | -0.91 | -1.58 to -0.24 | 0.007 |
| Family Caregiver Characteristics |  |  |  |
| Gender |  |  |  |
| Male (reference) | 0 |  |  |
| Female | -0.50 | -1.04 to 0.04 | 0.07 |
| Age | 0.03 | -0.01 to 0.06 | 0.10 |
| Relationship |  |  |  |
| Spouse (reference) | 0 |  |  |
| Child | -0.004 | -0.84 to 0.84 | 0.99 |
| Son-/daughter in law, Parent, Sibling, other | 0.99 | 0.27 to 1.71 | 0.007 |
| Education |  |  |  |
| Elementary school, high school (reference) | 0 |  |  |
| Vocational school, junior college, Undergraduate, Graduate | -0.25 | -0.74 to 0.23 | 0.31 |
| Presence of other caregivers |  |  |  |
| Present (reference) | 0 |  |  |
| Absent | -1.34 | -1.86 to -0.82 | p < 0.001 |
| Communication about the disease and life with the patient |  |  |  |
| Frequency (reference) | 0 |  |  |
| As needed | -2.09 | -2.56 to -1.61 | p < 0.001 |
| Rarely | -3.18 | -4.08 to -2.28 | p < 0.001 |

PCU: Palliative Care Unit

Adjusted R-squared, which is an indicator of effect size, was 0.12; the standard for effect size was moderate.

## Discussion

This nationwide survey, which was conducted in Japan, showed how congruence between a cancer patient’s preferred and actual place of death affected their quality of death and dying.

Our findings indicate that cancer patients who died in the place they preferred had better quality of life than patients who died in a place they did not prefer, as reported by their family caregivers.

This study used validated, comprehensive quality of death and dying measures specifically designed for terminally ill patients, and confirmed that congruence between preferred and actual place of death was strongly related to family-reported quality of death and dying. We found that quality of death and dying was higher in the achieved group than the not-achieved group. Moreover, the findings show that a better quality of death and dying was achieved in many components of good death beyond symptom relief (i.e., physical and psychological comfort), including spending time with family, maintaining hope and enjoyment, environmental comfort, and being respected as an individual. We believe that the total effect of these components may contribute to the achievement of better quality of death and dying.

Previous studies have shown that patients’ quality of death and dying was significantly higher at home relative to other places^6^. Our results were similar.

Those who died in acute-care hospitals and PCUs had GDI scores that were significantly higher for the achieved group compared with the not-achieved group. Nonsignificant differences were observed for those who died at home. This may be attributable to the smaller difference between the achieved and not-achieved groups in the GDI domains for the home setting than those for hospitals or PCUs. Several limitations in the present study should be noted. First, the participants were limited to those who had lost a loved one in a facility that was a member of HPCJ. Therefore, the findings may not represent the entire population of terminally ill cancer patients. Second, we could not rule out recall bias. According to previous surveys, 3 to 12 months after bereavement may be an appropriate time frame for participant inclusion, considering both recall bias and the grieving process [19–22]. Third, the nature of the study design did not allow us to evaluate the presence of pre-existing psychiatric disorders in family members during the period of patient death. Such disorders may affect the prevalence of depression and complicated grief during bereavement. Despite these limitations, the results of this study offer important insights and practical guidance for the period of time in which cancer patients and their families determine the place for end-of life care and place of death. Future research is needed to determine why a preferred place of death results in better end-of-life quality for patients.

### Conclusion

Congruence between preferred and actual place of death was associated with better quality of death and dying among terminally ill patients with cancer. This factor should be regarded as one of the essential components of end-of-life care in both clinical and political decision-making.

## Data Availability

All relevant data are within the manuscript and its Supporting Information files.

## Acknowledgments

We thank Anita Harman, PhD, from Edanz (https://jp.edanz.com/ac) for editing a draft of this manuscript.

## Supporting information

**S1 Fig. GDI Domain Scores for Death at Home.**

**S2 Fig. GDI Domain Scores for Death in an Acute-Care Hospital. S3 Fig. GDI Domain Scores for Death in a PCU.**

**S1 Table. GDI Scores for Congruence Between Preferred and Actual Place of Death Across Groups.** GDI: Good Death Inventory

